# Care dependency among older women in Austrian nursing homes: The role of outdoor temperatures

**DOI:** 10.1101/2020.09.07.20189563

**Authors:** Gerhilde Schuettengruber, Franziska Grossschaedl, Manuela Hoedl

## Abstract

The aim of this study was to investigate the associations between outdoor temperatures and care dependency degree among female nursing home residents. A secondary data analysis of data from a cluster-randomized controlled trial was conducted. Data from 345 female nursing home residents in two federal states in southern Austria were collected. Data on the degree of care dependency, dementia and age were collected from the participating female residents. Outdoor temperature data were provided by the Austrian Central Institute for Meteorology and Geodynamics. The results of the regression analysis show that the outdoor temperatures significantly affect the degree of care dependency. As the outdoor temperature decreases, the CDS score increases, which indicates that the number of independent woman enhance. For nursing home practice, we strongly recommend establishing heat plans. Representative prospective studies need to be carried out to investigate the causality between temperature and the degree of care dependency.

## Introduction

Climate change, a global phenomenon, represents one of the greatest environmental challenges of the twenty-first century^1^. Global warming, in terms of a climate change, seriously affects health and wellbeing^2^. A recent review reported that the number of articles that address the effects of climate change on health is still limited in comparison to the number of articles addressing these effects in other sectors, such as transportation and industry, indicating that further research on climate change and health is needed^3^. Although this area of research is rapidly growing^4^, especially quantitative studies^5^ that place a focus on the impact of climate change on health are still needed^6,7^. Heat-related health has been assigned an increasing level of priority all over the world^2^, and the WHO stated that research on climate change and health should include research on the most vulnerable groups^8^. Furthermore, in 2017, the Lancet Countdown’s highlighted the need to measure how vulnerable people were when they were exposed to climate change over time and investigate the influence of increasing temperatures on existing health problems^9^. In their recent review, Leyva, Beaman and Davidson (2017) reported that most studies on climate change and health have placed a focus on mortality rates related to cardiovascular diseases, while other studies have examined mortality related to respiratory diseases, hospitalization risk and increasing mental health problems^10^. As a consequence, estimates of approximately 1000 additional annual deaths of older people have been proposed due more frequent and intense heat waves for the period of 2036 –2065. When the chronically ill are included, a six-fold increase in the death rate in the older population is expected^11^.

Climate change and heat affect vulnerable populations to a greater extent than less vulnerable groups^9^. Older people are one major vulnerable population group with regard to heat and health issues^12^-^19^. As an international impact, the field of “climate gerontology,” has steadily developed over the past years^16^. Reported heat-related illnesses in older members of the population include heat cramps, heat exhaustion, heatstroke^12,20^, cerebro-/cardiovascular and respiratory outcomes^10^ and fluid/electrolyte disorders, renal failure, urinary tract infection and septicemia^20^.

One explanation for the increased vulnerability of older people to heat is that functional limitations also result in a reduction in their capacity to adapt to heat^12,13^. The homebound lifestyle of the older people as well as their lack of contact with other people are also factors that contribute to an increased risk of contracting a heat-related illness^12^. These aspects (pathophysiology, functional limitations and a homebound lifestyle) are especially applicable to nursing home residents. This statement echoes that made by Bittner and Stossel (2012), who argued that nursing home residents and people with dementia seem to be at the greatest risk during heat waves. The authors also highlighted that these high-risk groups also might be underrepresented in studies.

Overall, many studies have described the effects of heat on older people in terms of their mortality and morbidity. However, few studies have addressed specific, heat-related nursing phenomena, such as the degree of care dependency and the ability of older people to adapt to heat^10^.

Care dependency is defined as a “process in which the professional offers support to a patient whose self-care abilities have decreased and whose care demands make him/her to a certain degree dependent, with the aim of restoring this patient’s independence in performing self-care”^21^. The Care Dependency Scale (CDS) was developed to measure degrees of care dependency^21^. The scale comprises physical, psychosocial and spiritual aspects, and measurements taken using this scale allow caretakers to obtain a holistic view of care-dependent people.

In the nursing home setting, studies using this psychometrically tested scale have shown that more than half of nursing homes residents are completely or to a great extent care dependent^22,23^. While nursing home residents and people with dementia seem to be at the greatest risk in heat waves^17^, and many studies have described the effects of heat on older people in terms of their mortality and morbidity^10^, some study findings have shown that older women, in particular, are at greater risk of heat-related mortality than men^6,24,25^. However, no studies have placed a focus on the effects of different outdoor temperatures on the degree of care dependency of nursing home residents.

Therefore, the aim of this study was to investigate the association between outdoor temperatures and degree of care dependency in female nursing home residents in two federal states in southern Austria. We formulated the hypothesis that an increase in outdoor temperature decreases the number of completely care independent individuals among older female nursing home residents.

## Material and methods

### Design and Setting

In this study, a secondary data analysis was conducted based on data collected during a cluster randomized trial among Austrian female nursing home residents. A detailed description of the sample size calculation and other information can be found on the clinical trial registration website of the U.S. National Library of Medicine (no. NCT03030144). All nursing homes in the Austrian federal states of Styria and Carinthia that had ≥ 50 beds were invited by means of a letter and/or e-mail to participate in our study. Styria and Carinthia are located in the southern Austria.

### Sample

All female nursing home residents that were living in the participating nursing homes and who were assumed to stay there during the whole study period (3 months) were invited by the third author and a nurse to participate in the study. A sample size of 600 female residents was calculated. 676 female residents were invited, 295 of whom did not want to participate. Due to death, transfers to other institutions and other reasons, we included data from a total of 345 female residents in this analysis.

### Instrument used and data collection

The data collection was based on the Austrian questionnaire of the “International Prevalence Measurement of Care Problems” project^26^, including participation (yes/no), age, medical diagnoses of dementia or the nurses’ clinical assessment of cognitive impairment (yes/no) and the German version of the Care Dependency Scale (CDS)^21^. The CDS was developed by Dijkstra, Buist, and Dassen (1996) and is based on the fourteen basic human needs as described by Virginia Henderson. It is used to measure the degree of nursing care dependency. The scale comprises fifteen items, such as eating and drinking and continence. Each item of the CDS can be rated and assigned a score that ranges from one to five. One is assigned if the resident is completely dependent; two, if the resident is to a great extent dependent; three, in cases of partially dependency; four, in cases of limited extent dependency; and five, if the person is almost independent. The score given on the item level leads to a sum score over the whole scale, whereby higher scores indicate lower degree of care dependency (15-75)^21^. The German version of the CDS has been tested for its validity and reliability in nursing homes and hospitals^27,28^.

The third author and a nurse in each nursing home collected data at three measurement times that were convenient for the responsible nurse manager (baseline, six weeks, twelve weeks) between 19 April until 13 September 2017 in each nursing home. The baseline measurements taken in April and May were taken when the average outdoor temperature was 12°C, and the second measurements were taken between the end of May and the beginning of July, with an average outdoor temperature of 21°C. The third measurements were taken between mid-July and mid-September, with an average outdoor temperature of 20°C. The participating nursing homes did not have air conditioning in the residents’ rooms. Temperature data (measured in degrees Celsius) for the different regions of the participating nursing homes were provided by the Austrian Central Institute for Meteorology and Geodynamics (ZAMG).

### Analytic strategy

Analyses were conducted using SPSS Version 24.0^29^. We conducted descriptive statistics for sample characteristics, CDS variables and outdoor temperatures. The differences between the metric variables were calculated according to T-test. To test our hypothesis regarding the association between outdoor temperature and the assigned CDS, a logistic regression was calculated due to the fact that CDS is an ordinal scale. The independent effects of age (as a metric variable) and dementia (as a dichotomous variable), as possible covariates in the model, were also examined. Dementia was integrated into the model because the subjects concerned belong to a high-risk group during heat waves^17^. The model was chosen by goodness of fit scores in several steps on each measurement time point: (1) “Model Fitting information” with higher scores explaining a higher proportion of the variance of the dependent variable; (2) “Goodness of Fit” through Pearson (with scores of ≤ 0.05 indicating a good model); and (3) pseudo-R2 statistics with higher scores and tests for parallel lines of ≤ 0.05 indicating a good model. The model was tested such that the CDS represented a dependent variable for each measurement time point, starting with the average temperature, average temperature and age, average temperature and dementia, average temperature and age and dementia, as well as average temperature, age and dementia including the interaction of age and dementia. The category “completely independent” on the CDS was used as reference category in the model. Results are presented as parameter estimates, and 95% confidence intervals (95% CI) were calculated. Statistical significance was defined as *p* ≤ 0.05.

### Ethics

Two institutional review boards approved the study protocol (Styria: 29–007 ex 16/17; Carinthia: MZ 28/16) and written informed consent was obtained from all participating residents or their legal representatives.

## Results

Six nursing homes located in the Austrian federal states of Carinthia Styria, respectively, including a total of 345 female residents, were included in the analysis. The sample included participants who were 84 years old on average (SD = 10.7 years); the youngest woman was 33 and the oldest, 103 years old. Among the investigated women, 46.7% suffered from dementia. Table 1 shows the outdoor temperatures and CDS at the three measurement time points during the study period.

**Table 1.** Outdoor temperature and CDS scores during the study period

|  | **T1 (N = 345)** | **T2 (N = 345)** | **T3 (N = 345)** |
| --- | --- | --- | --- |
| Average outdoor temperature in °C* | 11.9 | 20.7 | 19.5 |
| Mean maximum outdoor temperature in °C* | 17.6 | 27.6 | 26 |
| Mean CDS sum score | 57.4 | 56.3 | 56.4 |
| **CDS categories** |  |  |  |
| Completely dependent (%) | 7.5 | 8.4 | 7.5 |
| To a great extent dependent (%) | 13.9 | 16.8 | 14.8 |
| Partially dependent (%) | 20.3 | 20 | 26.4 |
| To a great extent independent (%) | 25.2 | 21.2 | 22.0 |
| Almost independent (%) | 33 | 33.6 | 29.3 |

T 1 = Baseline, T2 = after 6 weeks, T3 = after 12 weeks; \* $p \leq 0.05$

The highest average temperature was measured at T2, and the highest CDS scores were found at T1, which means a lower grade of care dependency with regard to the sum score. The number of women who were completely care dependent or to a great extent care dependent was highest at T2. At all measurement time points, most of the residents were to a great extent independent or almost independent. The number of participants who were to a great extent independent or almost independent declined between T1 and T3. Figure 1 displays the number of participants that were almost independent for each CDS item, stratified by three measurement points.

**Fig. 1.**
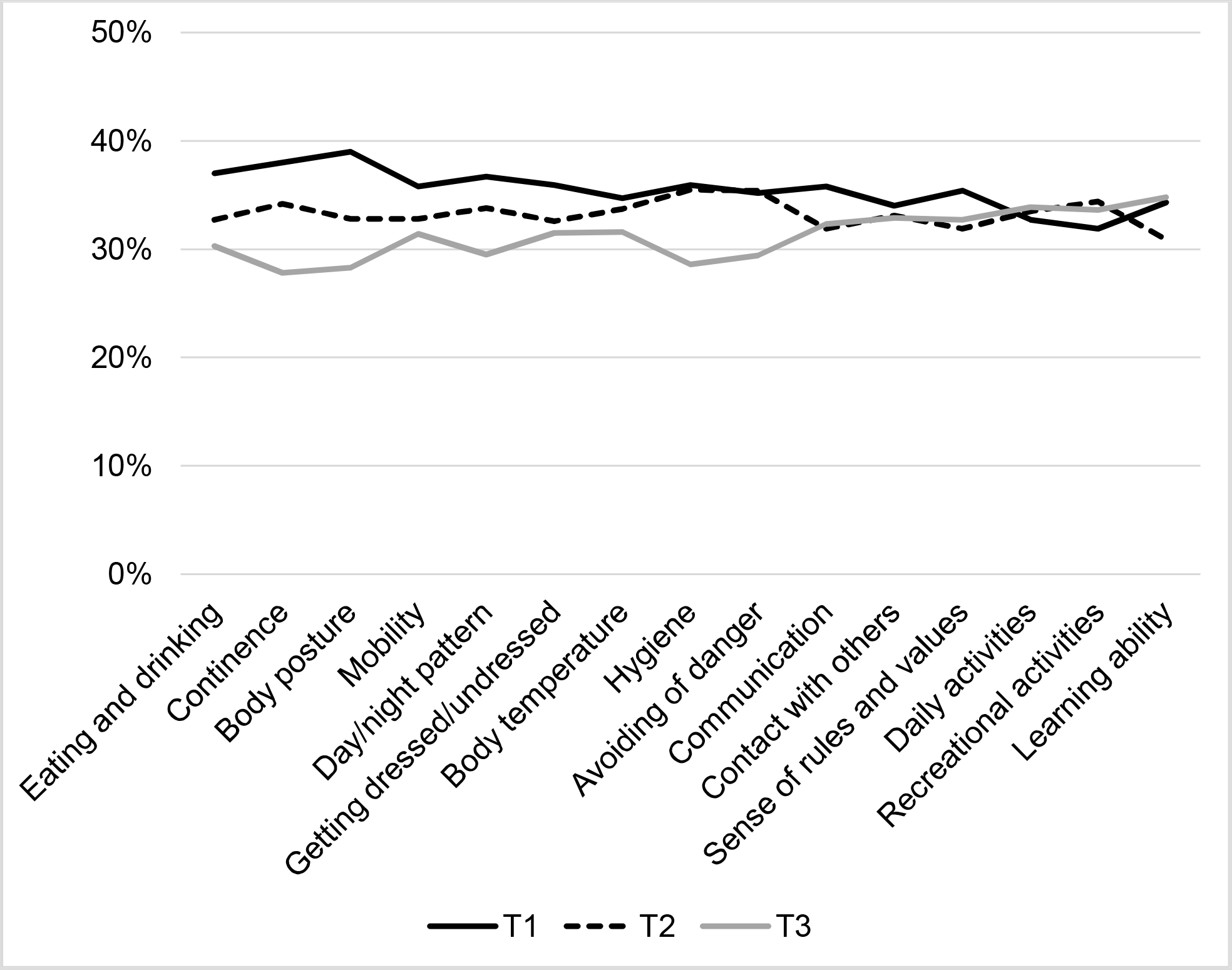
Number of participants that were almost independent for each CDS item, stratified by three measurement points (*N* = 345).

The results of the data analysis on the item level showed that the number of almost care independent women declined in all items, except daily activities, recreational activities and learning abilities. In the items eating and drinking, continence, body posture, mobility, day/night pattern, getting dressed/undressed, body temperature and hygiene, which can be labeled as functional items, the rate of decline observed was higher than that for the non-functional items (Fig. 1).

The results of the regression analysis (Tab. 2) over the three measurement points show that the outdoor temperature had a significant effect on the degree of care dependency. At T2, the time point with the highest temperature, the average and maximum outdoor temperature had a statistically significant impact on the degree of care dependency. As the outdoor temperature decreased, the CDS score increases, which indicates that the number of independent women increased. At T3, a slightly positive significant association between temperature and care dependency was observed. Dementia could be significantly associated with the degree of care dependency at all measurement time points. No significant associations were found for the item “age”.

**Table 2.**
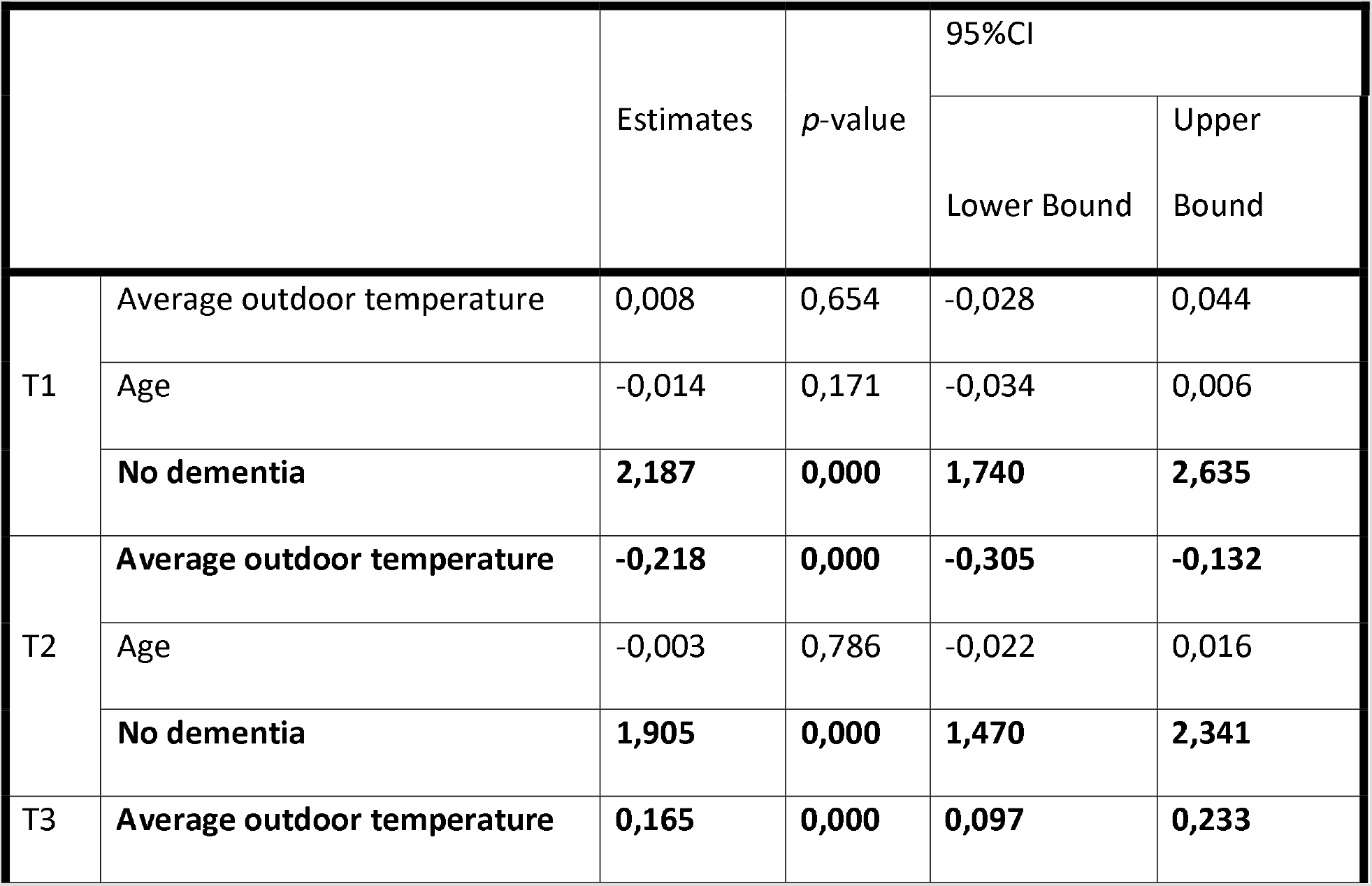
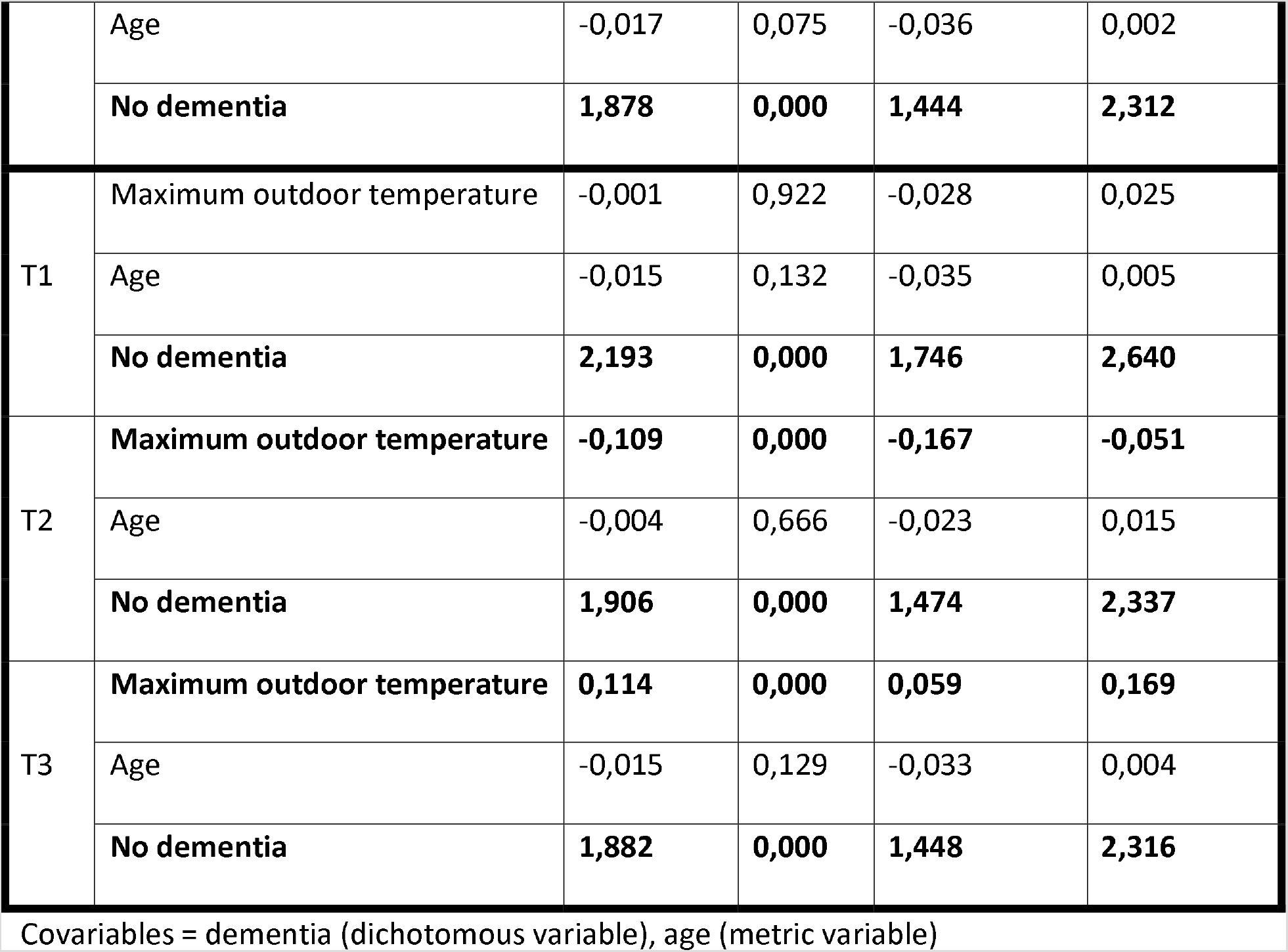
Results from the ordinal regression model on CDS for average and maximum outdoor temperatures, stratified by three measuring time points (*N* = 345)

## Discussion

Our results show that the outdoor temperature affected the degree of care dependency of women living in nursing homes in Styria and Carinthia. In addition, patients who suffered from dementia were normally had high degrees of care dependency at all measurement time points. Our results also showed that fewer female residents were almost care independent for CDS item levels at T3, especially regarding the functional items, than at T1 and T2.

Our findings showed that higher outdoor temperatures were positively associated with increases in the degree of care dependency among female nursing home residents. This can be explained by the fact that an increase in the number as well as the severity of health problems can lead to a higher degrees of care dependency. Such an increase in health problems has already been reported for heat-related illnesses in older people^10,20^.

Dementia was also significantly associated with a high degree of care dependency at all measurement time points independent of the outdoor temperature. This finding is in contrast to a conclusion from another study, in which qualitative interviews were conducted, where people with dementia seemed to be at most high risk during heat waves^17^. Due to the use of different methods, however, the results of these studies are not directly comparable.

Older people have been described in various studies as a major vulnerable group regarding heat and health issues^12-19^. Age was not a significant factor in our regression analysis. One reason for this might be that the sample in our study was quite homogenous regarding age; older women tended to participate.

The analysis on an item level mainly revealed a decline in functional items like eating and drinking, continence, body position, day and night pattern, hygiene and avoidance of danger. It is well known that those items, and especially continence, are also often affected by dementia, which was significantly associated with the degree of care dependency^30,31^.

Lindemann et al. (2018) demonstrated that increases in indoor temperature affected physical performance^32^. Eighty-one independent older adults with a mean age of 81 years (84% women) were investigated. The results of this study showed that high indoor temperatures affected the women’s physical capacity, chair-rise time, balance and gait speed. These findings support our hypothesis that the air temperature affects the functional abilities of older people and, therefore, their degree of care dependency. Another functional item affected by increasing temperature was the day-and night pattern. This result is in line with the results of an US survey study, where increased nighttime temperatures enhanced the number of self-reports of insufficient sleep, in general, as well as in the subsample of older adults^33^. A study that included 602 healthy participants reported that high air temperatures increased fatigue, especially for those with poor sleep patterns^34^. The findings of this study were also underpinned by those of a study in which the quality of sleep was correlated with changes in air temperature^35^.

In our study, the non-functional items, such as avoidance of danger, contact with others, sense of rules and values and learning ability, were also affected by outdoor temperatures. If we place a focus on the item “learning abilities,” another study on students showed a statistically significant decline in cognitive functions during heat waves, when no air conditioning was available in the building where they were living^36^. In another article, Lindemann (2018) showed that heat waves also have an effect on the social participation of older people. The author demonstrated an association between higher temperatures and declining social participations, especially among frail participants^32^. In contrast to our results, the latter study showed a slight increase in the number of independent residents as the heat increased with regard to their daily or recreational activities. These results could be explained by the use of different measurement instruments. Lindemann et al. (2018) asked questions that placed a focus on the general activity level, how time was normally spent, the amount of activity and participation in the community. The CDS used in this study placed a focus on daily activities, such as therapies or mealtimes and recreational activities like watching TV or reading.

Currently, a European working group is studying frailty related to heat waves, as a functional outcome, with the aim to prevent physical and functional declines in individuals during heat waves^37^. As frailty – in terms of physical and functional decline – can lead to increased care dependency, the work being carried out on heat-related frailty underpins the necessity of our analysis and highlights our results.

### Strengths and Limitations

One major strength of our study is the quality of the data obtained regarding to care dependency, which were collected by one of the authors together with a nurse in each participating nursing home. Another strength is that the association between care dependency and outdoor temperature could be investigated for the first time to our knowledge. This is also true with respect to the focus placed on female nursing home residents.

Our low sample size can be explained by the fact that, at baseline, 676 available female residents were invited for participation. Our sample size calculation was based on an assumed number of 950 female residents. The difference (676 vs. 950) can be explained by the fact that only two nursing homes participated with selected wards and some nursing homes were not fully booked. A larger sample would have resulted in more accurate outcomes and more meaningful models for regression analyses.

Another limitation was that indoor temperatures would have represented better and more precise variables, because they had the potentially to more strongly influence the results. Most primary studies use indoor temperature, but this was not possible in our study, since secondary data analysis was performed. For this reason, as well, men were not included.

## Conclusions

Heat should be considered as an influencing factor regarding the degree of care dependency in older people with a special focus on their functional capacity. For nursing home practice, we recommend establishing heat plans that include ward-specific schedules for airing the room and developing informative brochures for the residents with recommendations for clothing, how to prevent dehydration, daily activities that can be performed during heat waves and so on.

A higher outdoor temperature is positively correlated with a higher number of residents with a higher degree of care dependency. Therefore, nursing directors should consider the increase in the number of more highly care dependent residents in summer in their human resource planning and staff roster.

Nurses should regularly measure care dependency in nursing homes, especially during periods when high temperature variations are experienced, to be able to identify an increase in care dependency early on and to plan subsequent interventions. To measure the degree of care dependency, we can recommend the CDS. It is a valid and reliable instrument, which is easy to handle in practice.

Special attention should be given to residents who are almost care independent, since they seem to be affected most strongly by changes in temperatures. More research on a representative sample would be necessary to gain a greater insight into the topic and obtain outcomes that are more precise, as well as identify differences between women and men. Prospective studies should also be conducted to more carefully investigate the causality between temperature and the degree of care dependency.

## Data Availability

Due to leag issues data can not be made available.

